# Sustainable Health Innovation for Global Health Equity: Solar-Powered MRI for Affordable Healthcare in Resource-Limited Settings

**DOI:** 10.64898/2026.05.07.26352684

**Authors:** Michael Papasavva, Gebeyehu Begashaw Abate, Joseph Piper, Cynthia Kahari, Naume Tavengwa, Clever Mazhanga, Dzi Chidhanguro, Anabel Tsungai Mutero, Loyce Fadzai Musiiwa, Vincent Giampietro, Ricardo Twumasi, Petter Clemensson, Carly Bennallick, Sean Deoni, Chandiwana Nyachowe, Robert Ntozini, Steven C.R. Williams, Andrew J Prendergast, Niall J. Bourke

## Abstract

**Introduction:** Magnetic resonance imaging (MRI) is central to neurological care, yet access remains profoundly inequitable in low- and middle-income countries, especially in rural health facilities where high costs and fragile electricity supply limit services. Ultra-low-field (ULF) portable MRI offers a way to expand access, but deployment in weak-grid settings requires robust affordable power. We characterized the power needs of a 0.064T portable ULF MRI system and assessed the feasibility of a solar-powered MRI-capable facility in a rural Zimbabwean clinic, which we believe to be the first of its kind in the world.

**Methods:** We measured the power draw of an ultra-low-field MRI session from a portable photovoltaic (PV) battery kit in the UK, quantifying scan, standby and energy use. We then monitored a PV-battery micro-grid supplying a protected circuit at an MRI-capable clinic in Shurugwi, Zimbabwe. Inverter telemetry was used to derive PV generation, load, battery state of charge (SoC) and grid import for working days in October-November 2025, spanning the end of the dry season and onset of the rainy season.

**Results:** In the portable configuration, a 64-minute MRI session consumed approximately 0.21 kWh, with standby demand of approximately 1.44 kWh per 24 hours. In clinic, mean PV generation was 9.10 kWh (SD = 1.34) and load 6.91 kWh, with zero recorded grid import and minimum daily SoC typically greater than or equal to 60%, including during the early rainy season.

**Conclusion:** An affordable PV-battery micro-grid can reliably support ULF MRI and associated research power loads in a rural, weak-grid clinic, offering a reproducible blueprint to narrow diagnostic equity gaps in resource-limited settings.

## Introduction

Magnetic resonance imaging (MRI) is a cornerstone of modern neuroimaging, yet access remains profoundly unequal. In many low- and middle-income countries (LMICs), especially in rural regions, conventional MRI is scarce or centralized in distant tertiary hospitals. For example, there are only 1.12 MRI units per million population (pmp) in LMICs compared to 26.53 MRI units pmp in high-income countries (HICs) (Altaf et al., 2024). Even where scanners exist, services are constrained by high costs, shortages of trained personnel, and unreliable electricity (Liu et al., 2021; Rowand et al., 2025). These barriers translate into reduced access to care, high travel costs, and ultimately, substantial clinical risk for critically ill patients who may require imaging to guide time-sensitive decisions. These constraints also limit local research capacity, perpetuating underrepresentation of rural and LMIC populations.

Bridging this access gap requires imaging solutions that can operate under infrastructural constraints. Recent advances in ultra-low-field (ULF) MRI (< 0.1 T compared to standard strength of typically 3T), offer a practical route to reduce this access gap (Abate et al., 2024; Altaf et al., 2024; Mazurek et al., 2021). One of the most established examples is the Hyperfine Swoop®, a highly mobile (motorised), cart-based head-only MRI, scanner designed for bedside imaging in standard clinical spaces (see Figure 1). Its compact, self-shielded design allows safe deployment in routine clinical spaces without a built-in MRI suite. The system provides T1-weighted and T2-weighted imaging, fluid-attenuated inversion recovery (FLAIR), and diffusion-weighted imaging with apparent diffusion coefficient (ADC) maps (Shen et al., 2021). Crucially, it is built around a permanent magnet (64 millitesla or 0.064 T), avoiding the superconducting electromagnets and cryogenic cooling required by conventional MRI scanners. As a result, it runs from standard mains electricity with sub-kilowatt peak demand (approximately 0.9 kW – Swoop Device Specfications), making continuous operation feasible in settings with limited electrical infrastructure.

**Figure 1:**
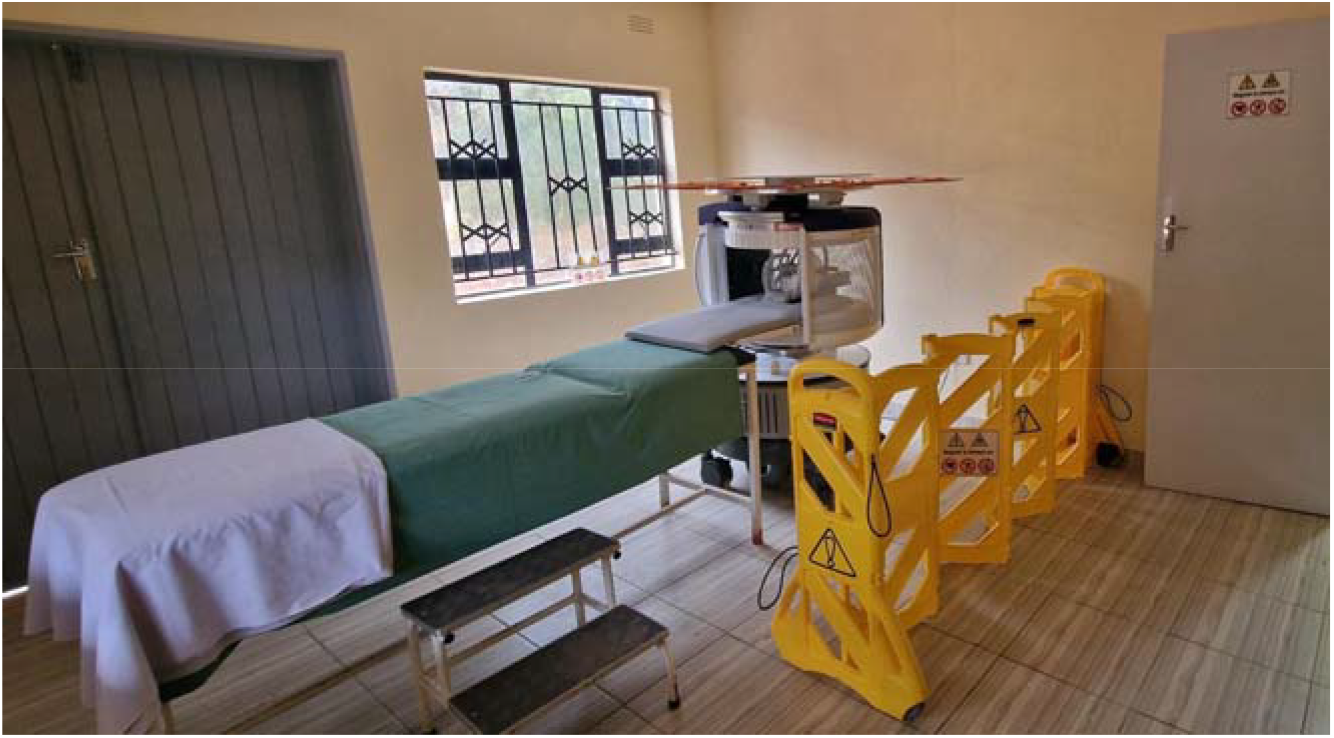
Swoop® Portable MR Imaging® System. Shurugwi Zimbabwe.

As ultra-low-field MRI is introduced across diverse clinical and research environments, standardisation becomes essential to ensure that imaging is comparable across sites and usable in practice. The Ultra-low-field Neuroimaging in The Young (UNITY) network, a multi-country consortium spanning more than 30 hospitals, universities, and clinical research centres, has supported deployment of ULF-MRI across LMICs by developing shared protocols for acquisition, quality control, training, and reporting (Abate et al., 2024). This has reduced operator variability, improved comparability of imaging data, and supported the growing use of ultra-low-field imaging in LMIC research (Cawley et al., 2023; Deoni et al., 2021). However, standardisation alone is insufficient. In many rural hospitals, unreliable electricity remains a major constraint: intermittent power, voltage fluctuation, and reliance on costly generator fuel can interrupt scans, disrupt data transfer, and damage ULF systems and associated networking infrastructure over time. In sub-Saharan Africa, about 15% of health-care facilities have no electricity access, and only 40% have reliable electricity (Organization, 2023). The challenge, therefore, is not only to introduce affordable imaging, but to sustain reliable service delivery in practice.

To address this, we integrated a Hyperfine Swoop® into a dedicated solar-powered micro-grid with battery storage at a custom-built research clinic housed in a government maternity hospital in rural Shurugwi (see Figure 3). To our knowledge, this is the first ever locally solar-powered deployment of an MRI machine globally. Consequently, the real-world performance and impact of solar-backed ULF MRI services in rural hospitals is currently absent from the medical and scientific literature. There is need to understand whether images can be acquired at all, and how often services might be available when needed. Evidence on operational metrics such as scan success and repeat rates, power-related interruptions, daily/seasonal variability, and upload reliability can inform deployment and scale-up decisions across UNITY and similar networks. Likewise, pragmatic descriptions of the power subsystem, from protective devices to cable sizing and remote telemetry, are essential for replication by research and government partners, and for hospital engineering teams.

Here, we report two complementary strands of work. First, we characterise the power requirements of the Hyperfine Swoop® under controlled conditions by operating the scanner from a portable photovoltaic (PV) and battery kit in the United Kingdom, isolating the MRI load from other clinical, research, and facility demands (Figure 2). This configuration provides scanner-specific power profiles that can be used to size renewable systems and to benchmark performance independently of local infrastructure. Second, we describe the design, implementation, and early research utility of a solar-powered, UNITY-affiliated MRI-capable research laboratory housing a separate Hyperfine Swoop®, located on the grounds of a rural district maternity hospital in Shurugwi, Zimbabwe. We include the site context, power architecture, and operational stability. By detailing both the controlled solar-powered experiment and the subsequent real-world rural deployment, we aim to provide scanner-level power metrics that can inform the design of larger solarised facilities where MRI will typically be one of several pieces of equipment and to create a reproducible blueprint for integrating ULF MRI into facilities in resource-constrained settings.

**Figure 2:**
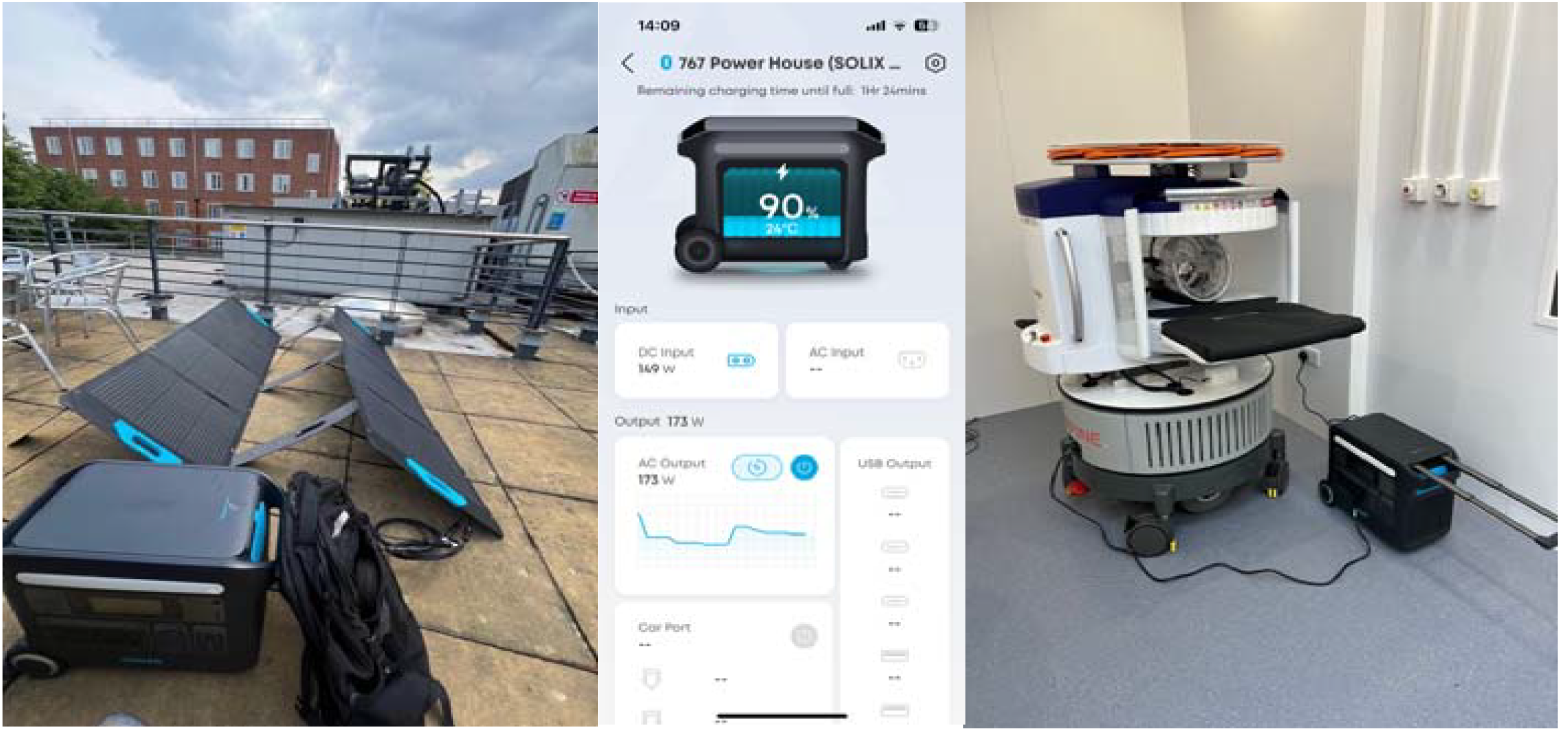
Anker Solix F2000 Portable Power Station pictured and Hyperfine Swoop. Kings College London.

## Methods

Ethical approval of the current work falls under the study for the Process and economic Evaluation of portable Ultra-Low Field MRI Technology Adoption in Low- and Middle-Income Countries (LMICs) healthcare settings (LRS/RGO-24/25-47006).

### Hyperfine Swoop power draw tests

Evaluation of a minimal low-cost power setup and power draw test on the Hyperfine Swoop® was performed at King’s College London. The Anker Solix F2000 Portable Power Station with 2.05 kWh usable capacity with 800w (4 × 200w) PV panels was tested as a commercially available portable low-cost setup to accompany the ULF scanner (Figure 2). This power-station charge can be supplemented by grid energy where available. Power draw tests were performed using a commercially available MECHEER power meter plug. KWh were recorded for each scanning sequence, and the cumulative power draw was recorded for a full 64min scanning session.

### Rural field site in Zimbabwe

To provide stable, continuous power for bedside imaging and data transfer, we designed a small photovoltaic (PV) and battery micro-grid supplying a protected clinical circuit. The architecture comprised two 5 kW Sunsynk hybrid inverters operating in parallel (230 V alternating current, 50 Hz) with remote monitoring; three 5.12 kWh lithium-iron-phosphate (LiFePO□) battery modules (15.36 kWh nominal capacity); and twelve 600 W monocrystalline PV modules (7.2 kWp total), mounted on IBR roofing with galvanised rails, IBR brackets, and anti-theft clips (Figure 3). Protection and wiring followed standard practice: an upstream 63 A alternating-current (AC) breaker; a 500 V / 20 kA AC surge-protection device; 20 A branch breakers; 6 mm^2^ PV direct-current (DC) cabling (red/black) and 6 mm^2^ four-core AC cabling to a dedicated sub-board; and a 10 mm^2^ earth conductor bonded to a 1.2 m galvanised earth rod. Critical loads on the protected bus were the MRI system, an online uninterruptible power supply (UPS) with voltage conditioning, the network router/access point, and essential lighting and ventilation.

**Figure 3:**
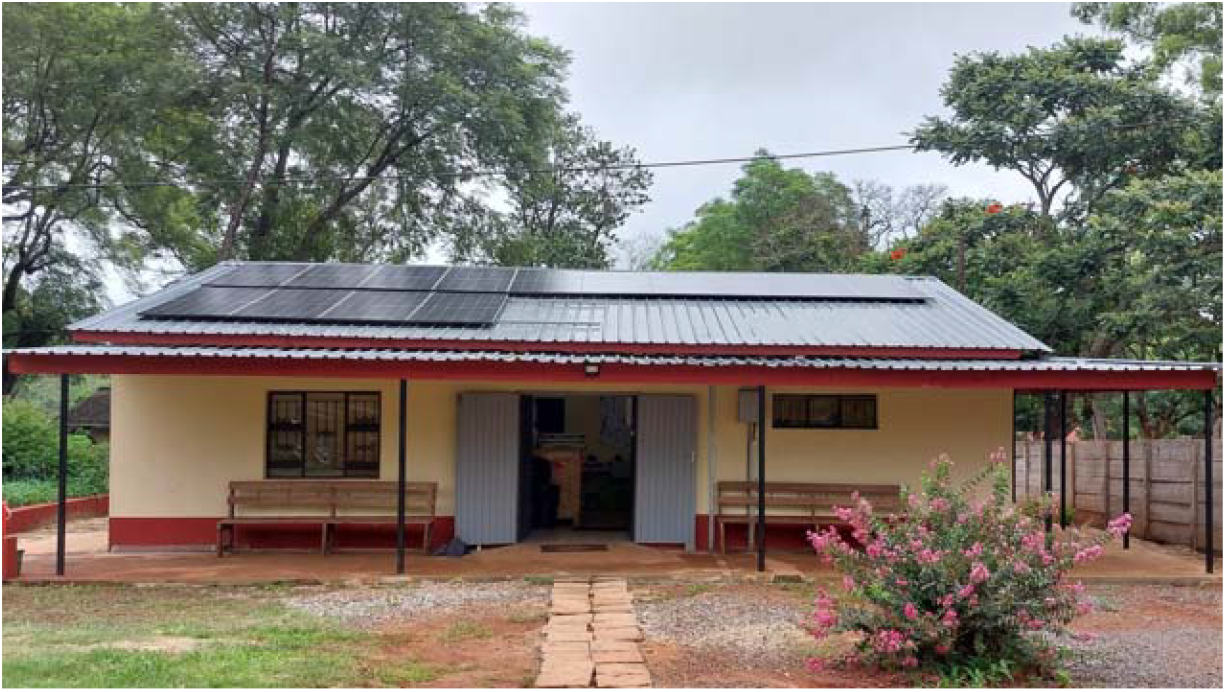
Zvitambo Clinical Research Facility. Shurugwi, Zimbabwe

**Figure 4:**
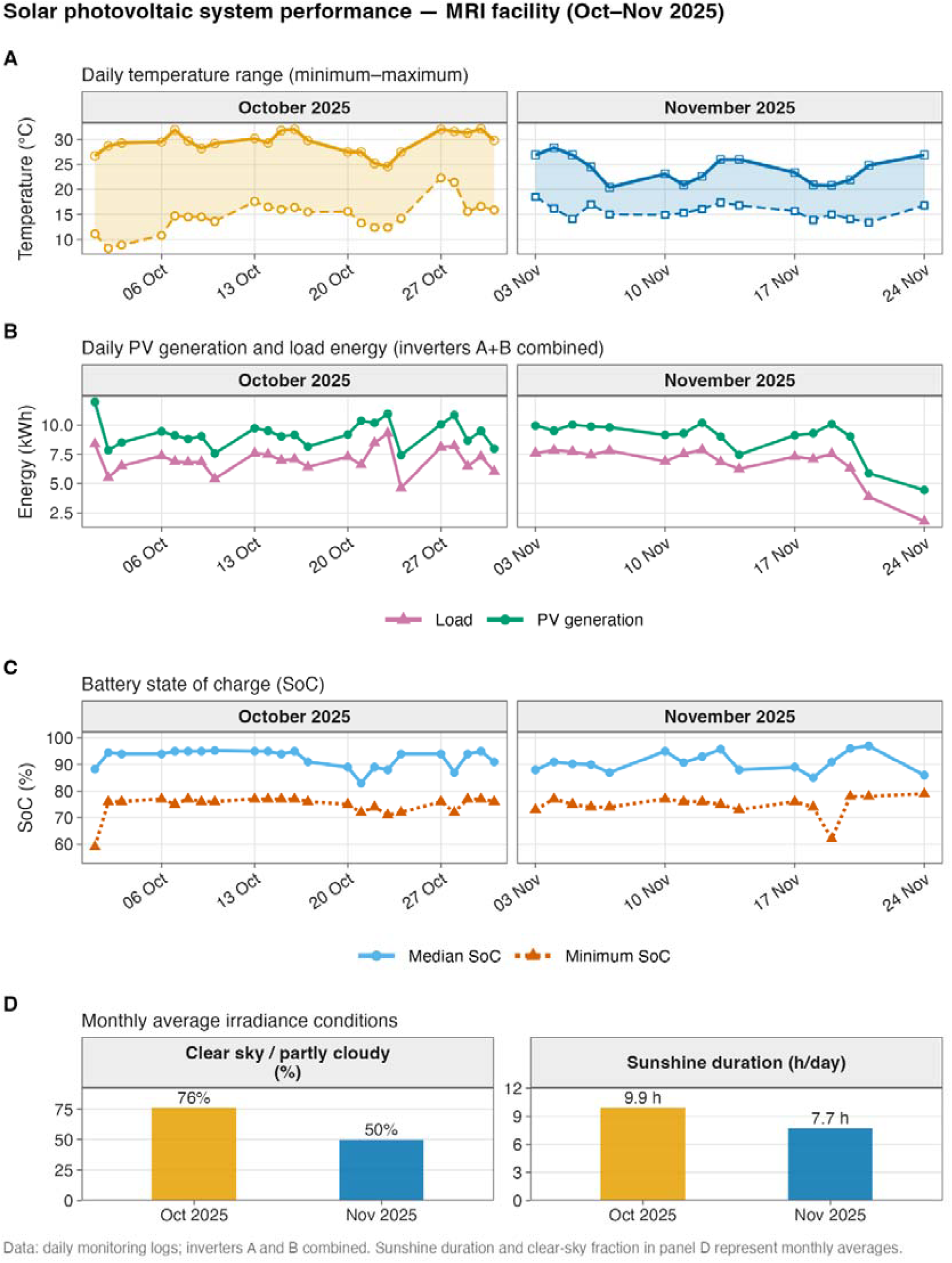
Shurugwi solar photovoltaic system performance

Grid sizing was based on the scanner’s measured/quoted demand (approximately 0.9 kW at acquisition peaks), and common clinical research equipment, including ultrasound, electroencephalography (EEG), peripheral quantitative computed tomography (pQCT), spirometry, and multiple laptops alongside lighting and networking requirements. At full utilisation, the Shurugwi site handled up to four participants per day, typically scheduled as two morning and two afternoon scans. For each participant we acquired a focused low-field MRI protocol in order to calculate brain volume estimates comprising multi-planar T2-weighted imaging in axial, coronal and sagittal orientations. In addition, a T1-weighted sequence optimised for grey–white matter contrast was acquired. This use provides interpretable data while keeping total scan time per participant sufficiently short to be compatible with the practical constraints of a rural research setting, particularly when scanning infants.

In Shurugwi, the public electricity grid is highly irregular, with frequent outages and voltage fluctuations, so the solar–battery system was designed to support routine operations largely independent of grid supply, with the grid treated as a secondary backup. The 8 kW inverter cluster provides transient headroom for short surges. Battery storage of 15.36 kWh nominal, approximately 12.3 kWh usable at 80% depth of discharge, supports around 7–12 hours of operation depending on ancillary loading and conversion losses. The 7.2 kWp PV array was sized to support daytime operation of an eight-room clinic (housing ultrasound, pQCT, EEG and laptop use) and to achieve typical overnight state-of-charge (SoC) targets.

In central Zimbabwe, October corresponds to the late dry season and is typically one of the hottest, sunniest months, with long sunshine duration and predominantly clear or partly cloudy skies. By contrast, November marks the onset of the rainy season, with lower average sunshine hours, greater cloud cover, and more day-to-day variability in irradiance, providing a more challenging operating period for the solar-backed MRI and laboratory loads. Performance across October and November 2025 were summarised, investigating working days using monthly averages of weather, PV generation, load energy, battery SoC, and grid import.

## Procedures

Operational telemetry was obtained using the Sunsynk monitoring portal, which reports inverter output, PV input, battery state of charge (SoC) and grid exchange. We characterised solar generation, load consumption and battery performance using log files from the two hybrid inverters supplying the MRI and laboratory circuits. Each inverter recorded time-stamped data at approximately 5-min intervals, including photovoltaic (PV) input power (ppv1–3), alternating-current (AC) load power at the uninterruptible power supply (UPS) / protected-load output (upsTotalLoad / loadTotalPower), battery state of charge (SoC; battery Energy), daily grid-import energy counter (gridBuyToday), and cumulative PV and load energy counters (pvetoday and dailyUsed / etoday). For each inverter and day, we converted the on-board time field to a POSIX (Unix) timestamp and estimated the median sampling interval (Δt, hours) from successive time differences. Instantaneous PV power was calculated as the sum of all available PV input channels, and instantaneous load power as the UPS/protected-load output. Daily PV and load energies were then obtained by numerical integration of these series over time (Σ power × Δt), with conversion from W to kWh.

Battery performance was summarised as the minimum and median SoC (%) observed each day. Daily grid import was calculated from the change in the cumulative grid-import counter (gridBuyToday) and aggregated across both inverters. For each working day with valid telemetry in October and November 2025, system-level PV and load energies were derived by summing across inverters, and system-level SoC by taking the lower of the two daily minima and the median of the two daily medians. As a validation step, integrated PV and load energies were compared against the cumulative counters (pvetoday for PV generation and dailyUsed / etoday for load consumption), with agreement typically within ∼0.1–0.5 kWh per day; we therefore used the integrated time-series values for analysis and treated the counters as a cross-check. Power analyses were restricted to working days in October and November 2025 when the laboratory was open and the facility was in routine use. Days when the facility was closed or no research activity was scheduled (including 25–28 November) were excluded a priori, even if limited telemetry was available.

## Results

### Hyperfine Swoop Power Draw

To assess the potential of a solar-powered setup for the scanner we measured the power draw on the Hyperfine Swoop®. We measured a draw of 0.21KWh for a 64-minute session (1.26 kWh/6hrs), including stock sequences and two advanced research sequences (T2 mapping, MT, DWI). Under standby conditions the Swoop® drew 0.06kWh (1.44kWh/24hrs).

**Table 1:** Hyperfine Swoop Sequence List Load Summary.

| **Sequence** | **KWh** |
| --- | --- |
| localiser | 0.005 |
| T2w axi fast | 0.013 |
| T2w sag fast | 0.011 |
| T2w cor fast | 0.01 |
| T1w contrast | 0.021 |
| T1w axi | 0.018 |
| MT | 0.019 |
| T2 mapping | 0.04 |
| DWI | 0.073 |

To explore the possibility of a low-cost minimal requirement set up we tested the Anker Solix F2000 Portable Power Station. During cloudy summer conditions in London 150Wh was generated from 400W solar portable solar panels (mean Temp = 24**°**C; 37.5% efficiency).

### Solar-powered rural site

Across October 2025 working days with valid telemetry (n = 23), corresponding to the end of the dry season in central Zimbabwe, mean daily minimum and maximum temperatures were M = 14.7 °C and M = 29.4 °C, respectively, with an average of M = 9.9 h of sunshine per day and clear or partly cloudy conditions for approximately 76% of the daytime period. Mean daily photovoltaic generation was M = 9.26 kWh (inverters A+B combined), with a corresponding mean daily load of M = 7.03 kWh. Grid import was 0.00 kWh on all observed days, and the battery operated at a mean minimum state of charge (SoC) of M = 74.7% and a mean median SoC of M = 92.4%. See figure four for a day-by-day summary of PV generation, load consumption, grid import and battery SoC statistics for October 2025.

For November 2025 working days with valid telemetry (n = 16), marking the onset of the rainy season, nights were slightly warmer and days cooler on average (Tmin M = 15.6 °C; Tmax M = 24.0 °C), with reduced sunshine (M = 7.7 h/day) and clear or partly cloudy conditions for approximately 50% of the daytime. Mean daily photovoltaic generation was M = 8.88 kWh and mean daily load was M = 6.73 kWh, again with zero recorded grid import. Despite lower irradiance and greater cloud cover than in October, battery reserves remained stable, with a mean minimum SoC of M = 74.8% and a mean median SoC of M = 90.8%. See Table 2 for a day-by-day summary of PV generation, load consumption, grid import and battery SoC statistics for November 2025.

Across both months, mean daily photovoltaic generation was approximately M = 9.1 kWh and mean daily load was M = 6.91 kWh, with no grid import recorded. On average, the site experienced approximately M ≈ 9.0 h of sunshine per working day and clear or partly cloudy conditions for roughly two-thirds of the daytime period, while the battery bank maintained a mean minimum SoC of around M = 74.7% and a mean median SoC of approximately M = 91.7%, indicating substantial reserve capacity even under cloudier, early–rainy-season conditions.

To set up this solar powered experiment in rural Zimbabwe maternal healthcare facility, a total solar cost of $11,810.80 was required. Over 50 % of this cost was spent to purchase main equipment, including the solar panel, inverter, and battery (see supplementary table 1).

## Discussion

We show that the power draw from an ultra-low-field (ULF) portable MRI system is sufficiently low to run off battery power stations and can be reliably powered using modest photovoltaic (PV)–battery configurations in a rural district hospital in Shurugwi, Zimbabwe. By combining scanner-level power characterisation from two scanners in London and inverter telemetry from a solar-backed clinic in Shurugwi, Zimbabwe, we provide quantitative evidence that a PV–battery micro-grid can support ULF MRI and associated research loads with no reliance on grid electricity across contrasting seasonal conditions.

First, we quantified the intrinsic power requirements of the Swoop® ULF MRI scanner: a typical 64-minute session consumed ∼0.21 kWh, while 24-hour standby drew ∼1.44 kWh. A major advantage of the ULF system is that it can be switched off entirely when not in use, providing additional energy savings by not having a continual standby power draw. These demands are minimal compared with conventional high-field MRI, which can require continuous high power and specialised electrical infrastructure. 3T scanners can exceed 12,000 kWh per month (average load 16-17 kWh) even when in standby for ∼75% of the time (Carver et al., 2026).

Adoption decisions in low-resource settings are often shaped by perceived rather than measured power needs; our data indicate that appropriately specified PV–battery systems can accommodate routine scanning, especially with basic load management (e.g., clustering scans during daylight and reducing unnecessary standby). Second, we assessed real-world performance in a rural facility with multiple clinical research loads and unreliable grid supply. Across working days in October–November 2025, the Shurugwi micro-grid met combined MRI and laboratory demand (mean daily load ∼7 kWh in October and ∼6.7 kWh in November) with zero recorded grid import. Minimum daily battery state of charge typically remained above 60%, indicating substantial reserve capacity even as the rainy season began. In practice, this represents genuine decoupling from grid instability rather than intermittent solar assistance.

These findings speak directly to diagnostic inequity. MRI access remains highly unequal, with limited availability in many low- and middle-income countries and concentration of scanners in urban centres. Our results suggest reliable MRI services need not be limited to sites with strong grids or large generator budgets: renewable-backed, decentralised imaging can be placed closer to patients and to cohort study sites, reducing travel and out-of-pocket expenditure, strengthening rural research capacity, and improving resilience to fuel shortages, price shocks, and grid failures while reducing dependence on diesel generators.

The Shurugwi system (7.2 kWp PV; 15.36 kWh) was designed to support an eight-room research clinic with additional equipment (e.g., ultrasound, EEG, pQCT), not just an isolated scanner. Despite these loads, daily PV generation (∼9 kWh in October and ∼8.9 kWh in November) exceeded or closely matched consumption, and the battery rarely approached deep discharge thresholds, suggesting ULF MRI can often be integrated without disproportionate oversizing. High-resolution inverter telemetry proved valuable for monitoring, identifying risk periods, and supporting optimisation.

Although this study focused on feasibility, it is worth noting that the upfront capital expense of the full PV–battery system was ∼USD 11,800 (including hardware, protection, logistics, and installation). When amortised and shared across multiple functions, this may compare favourably to recurring diesel costs, generator maintenance, and downtime from grid instability. We did not conduct a full cost-effectiveness analysis; future work should model cost per scan under different utilisation scenarios, incorporating life-cycle costs (maintenance and battery replacement) and potential savings from reduced generator use and avoided patient transfers. Throughput will be important: the Shurugwi site was configured for up to four research participants per day, and higher appropriate throughput would improve the economic case. The portable power station $3,000 USD is a smaller lower cost alternative, for lower demand use or backups where there is unreliable grid infrastructure. The portable power stations can also be used as an offline uninterrupted power supply. A full charge, assuming 75% efficiency, would have the potential for running the scanner for 6 hours.

This work is strengthened by pairing controlled bench testing with real-world deployment at an active UNITY site and by using high-frequency telemetry to provide additional quantitative evidence. It should be noted that, as a case-study, there is the limitation that reporting is from a single high-insolation setting; broader generalisability will require modelling and additional deployments in a range of insolation settings. We focused on power availability and did not systematically assess image quality, diagnostic impact, or clinical pathways. We also did not formally quantify grid outage frequency during monitoring, limiting counterfactual estimates of disruption without solar backing, and we do not yet provide full life-cycle costing. Our evaluation was limited to two calendar months and year-round performance needs to be ascertained.

Next steps include optimising PV–battery sizing for different climates and load profiles, including adding MRI to existing solarised clinics. Linking power telemetry with clinical operations (scan counts, cancellations, referrals) would enable more direct assessment of effects on access, timeliness, and outcomes. More broadly, pairing portable ULF MRI with locally owned solar micro-grids could extend to other imaging needs where electricity is a constraint, including trauma care, obstetrics, and oncology.

In summary, we have shown that a modest PV–battery micro-grid can reliably sustain ULF MRI and associated research activities in a rural, weak-grid hospital with ample reserve capacity across dry-season and early rainy-season conditions. Combined with scanner-level power characterisation, these findings provide a practical blueprint for solar-powered MRI in resource-limited settings, supporting diagnostic equity while reducing dependence on unstable grids and fossil fuels.

## Data Availability

All data produced in the present work are contained in the manuscript

## Supplementary materials

**Supplementary table 1:** Total Solar Costs for MRI Laboratory.

| Item | Description | Unit | Qty | Unit price (USD) | Total price (USD) |
| --- | --- | --- | --- | --- | --- |
| **MAIN EQUIPMENT** |  |  |  |  |  |
| 1 | Sunsynk inverter 5kW with remote monitoring function | No. | 2 | 1,001.00 | 2,002.00 |
| 2 | Sunsynk 5.12kWh 51.2V Battery | No. | 3 | 1,120.60 | 3,361.80 |
| 3 | 600W Jinko Monocrystalline Solar panel | No. | 14 | 119.60 | 1,674.40 |
| **MOUNTING STRUCTURE** |  |  |  |  |  |
| 4 | Galvanised Rails | No. | 8 | 24.70 | 197.60 |
| 5 | Anti-theft clips | No. | 72 | 0.98 | 70.20 |
| 6 | IBR Brackets | No. | 40 | 1.95 | 78.00 |
| 7 | Accessories (bolt & nuts) | lot | 1 | 75.00 | 75.00 |
| **EARTHING SYSTEM** |  |  |  |  |  |
| 8 | Earthing cable 10mm single core yellow/green | m | 30 | 2.09 | 62.79 |
| 9 | Earth rod galvanised 1.2m × 16mm 4-foot c/w clamp | No. | 1 | 30.71 | 30.71 |
| 10 | Accessories (lugs, earth clips etc.) | lot | 1 | 65.00 | 65.00 |
| **CABLING** |  |  |  |  |  |
| 11 | 6mm PV Solar cable Red to run 4 strings (approx. 30 m) | m | 120 | 1.43 | 171.60 |
| 12 | 6mm PV Solar cable Black to run 4 strings (approx. 30 m) | m | 120 | 1.43 | 171.60 |
| 13 | 6mm 4 Core AC Cable | m | 25 | 8.06 | 201.50 |
| 14 | 50 × 50 mm Trunking | No. | 4 | 9.75 | 39.00 |
| 15 | Accessories (cable ties, insulation tape, MC4 connectors, DC Bus Bar chamber, etc.) | lot | 1 | 97.50 | 97.50 |
| 16 | 35mm Battery Cable Red and black | No. | 6 | 10.40 | 62.40 |
| **PROTECTION SYSTEM** |  |  |  |  |  |
| 17 | AC SPD 500V 20KA | No. | 2 | 19.50 | 39.00 |
| 18 | AC 20A Breaker | No. | 4 | 2.59 | 10.35 |
| 19 | 63A AC Breaker | No. | 2 | 2.78 | 5.56 |
| 20 | 40A AC Breaker | No. | 2 | 2.99 | 5.98 |
| 21 | AC Combiner box | No. | 1 | 19.50 | 19.50 |
| 22 | 63A AVS | No. | 1 | 19.50 | 19.50 |
| 23 | 63A Changeover switch | No. | 1 | 9.10 | 9.10 |
| 24 | 600V DC SPD | No. | 4 | 19.50 | 78.00 |
| 25 | 20A DC Breaker DP | No. | 4 | 5.20 | 20.80 |
| 26 | DC combiner box | No. | 1 | 19.50 | 19.50 |
| **ADMINISTRATION COSTS** |  |  |  |  |  |
| 27 | Transport and logistics | lot | 1 | 945.00 | 945.00 |
| 28 | Training | days | 1 | 175.00 | 175.00 |
| 29 | Installation & commissioning | days | 2 | 230.00 | 460.00 |
| 30 | Ps & Gs' | lot | 1 | 79.46 | 79.46 |
|  | SUB TOTAL |  |  |  | 10,270.26 |
|  | VAT (15%) |  |  |  | 1,540.54 |
|  | TOTAL PAYABLE IN USD |  |  |  | 11,810.80 |

**Supplementary Table 2:** October 2025 Working Days – Shurugwi Weather, Solar and Load Summary.

| Date | Day | Tmin<br>(°C) | Tmax<br>(°C) | Sunshine<br>hours<br>(h/day)<br>Monthly<br>average | Clear<br>sky /<br>partly<br>cloudy<br>time<br>(%)<br>average | Photovoltaic<br>energy (kWh,<br>Inverters<br>A+B) | Load<br>energy<br>(kWh,<br>Inverters<br>A+B) | Battery<br>SoC<br>(min /<br>median,<br>%) | Grid<br>energy<br>bought<br>(kWh,<br>Inverters<br>A+B) |
| --- | --- | --- | --- | --- | --- | --- | --- | --- | --- |
| 2025-10-01 | Wed | 11.1 | 26.7 | 9.9 | 76 | 11.97 | 8.38 | 59 /<br>88.25 | 0.00 |
| 2025-10-02 | Thu | 8.2 | 28.7 | 9.9 | 76 | 7.84 | 5.52 | 76 /<br>94.50 | 0.00 |
| 2025-10-03 | Fri | 8.9 | 29.3 | 9.9 | 76 | 8.50 | 6.50 | 76 /<br>94.00 | 0.00 |
| 2025-10-06 | Mon | 10.8 | 29.5 | 9.9 | 76 | 9.45 | 7.37 | 77 /<br>94.00 | 0.00 |
| 2025-10-07 | Tue | 14.7 | 31.9 | 9.9 | 76 | 9.11 | 6.88 | 75 /<br>95.00 | 0.00 |
| 2025-10-08 | Wed | 14.5 | 29.7 | 9.9 | 76 | 8.81 | 6.82 | 77 /<br>95.00 | 0.00 |
| 2025-10-09 | Thu | 14.5 | 28.2 | 9.9 | 76 | 9.05 | 6.85 | 76 /<br>95.00 | 0.00 |
| 2025-10-10 | Fri | 13.6 | 29.2 | 9.9 | 76 | 7.56 | 5.40 | 76 /<br>95.25 | 0.00 |
| 2025-10-13 | Mon | 17.6 | 30.2 | 9.9 | 76 | 9.73 | 7.59 | 77 /<br>95.00 | 0.00 |
| 2025-10-14 | Tue | 16.5 | 29.3 | 9.9 | 76 | 9.51 | 7.49 | 77 /<br>95.00 | 0.00 |
| 2025-10-15 | Wed | 16.0 | 31.8 | 9.9 | 76 | 9.03 | 6.98 | 77 /<br>94.00 | 0.00 |
| 2025-10-16 | Thu | 16.4 | 32.0 | 9.9 | 76 | 9.15 | 7.09 | 77 /<br>95.00 | 0.00 |
| 2025-10-17 | Fri | 15.5 | 29.8 | 9.9 | 76 | 8.15 | 6.41 | 76 /<br>91.00 | 0.00 |
| 2025-10-20 | Mon | 15.6 | 27.5 | 9.9 | 76 | 9.18 | 7.29 | 75 /<br>89.00 | 0.00 |
| 2025-10-21 | Tue | 13.3 | 27.5 | 9.9 | 76 | 10.38 | 6.60 | 72 /<br>83.00 | 0.00 |
| 2025-10-22 | Wed | 12.4 | 25.2 | 9.9 | 76 | 10.17 | 8.47 | 74 /<br>89.00 | 0.00 |
| 2025-10-23 | Thu | 12.4 | 24.6 | 9.9 | 76 | 10.95 | 9.28 | 71 /<br>88.00 | 0.00 |
| 2025-10-24 | Fri | 14.2 | 27.5 | 9.9 | 76 | 7.43 | 4.63 | 72 /<br>94.00 | 0.00 |
| 2025-10-27 | Mon | 22.3 | 32.0 | 9.9 | 76 | 10.05 | 8.10 | 76 /<br>94.00 | 0.00 |
| 2025-10-28 | Tue | 21.4 | 31.6 | 9.9 | 76 | 10.84 | 8.22 | 72 /<br>87.00 | 0.00 |
| 2025-10-29 | Wed | 15.6 | 31.3 | 9.9 | 76 | 8.67 | 6.47 | 77 /<br>94.00 | 0.00 |
| 2025-10-30 | Thu | 16.6 | 32.1 | 9.9 | 76 | 9.49 | 7.30 | 77 /<br>95.00 | 0.00 |
| 2025-10-31 | Fri | 15.9 | 29.8 | 9.9 | 76 | 7.98 | 6.03 | 76 /<br>91.00 | 0.00 |

**Supplementary Table 3:** November 2025 Working Days – Shurugwi Weather, Solar and Load Summary.

| Date | Day | Tmin (°C) | Tmax (°C) | Sunshine hours (h/day) | Clear sky / partly cloudy time (%) | Photovoltaic energy (kWh, Inverters A+B) | Load energy (kWh, Inverters A+B) | Battery SoC (min / median, %) | Grid energy bought (kWh, Inverters A+B) |
| --- | --- | --- | --- | --- | --- | --- | --- | --- | --- |
| 2025-11-03 | Mon | 18.5 | 26.9 | 7.7 | 50 | 9.94 | 7.60 | 73 / 88.00 | 0.00 |
| 2025-11-04 | Tue | 16.2 | 28.3 | 7.7 | 50 | 9.51 | 7.84 | 77 / 91.00 | 0.00 |
| 2025-11-05 | Wed | 14.1 | 26.9 | 7.7 | 50 | 10.02 | 7.72 | 75 / 90.25 | 0.00 |
| 2025-11-06 | Thu | 17.0 | 24.5 | 7.7 | 50 | 9.87 | 7.44 | 74 / 90.00 | 0.00 |
| 2025-11-07 | Fri | 15.0 | 20.4 | 7.7 | 50 | 9.79 | 7.81 | 74 / 87.00 | 0.00 |
| 2025-11-10 | Mon | 14.9 | 23.1 | 7.7 | 50 | 9.15 | 6.89 | 77 / 95.00 | 0.00 |
| 2025-11-11 | Tue | 15.3 | 20.9 | 7.7 | 50 | 9.28 | 7.53 | 76 / 90.75 | 0.00 |
| 2025-11-12 | Wed | 16.1 | 22.6 | 7.7 | 50 | 10.17 | 7.88 | 76 / 93.00 | 0.00 |
| 2025-11-13 | Thu | 17.4 | 26.0 | 7.7 | 50 | 9.02 | 6.84 | 75 / 95.75 | 0.00 |
| 2025-11-14 | Fri | 16.8 | 26.0 | 7.7 | 50 | 7.46 | 6.24 | 73 / 88.00 | 0.00 |
| 2025-11-17 | Mon | 15.7 | 23.4 | 7.7 | 50 | 9.12 | 7.30 | 76 / 89.00 | 0.00 |
| 2025-11-18 | Tue | 13.9 | 20.9 | 7.7 | 50 | 9.29 | 7.09 | 74 / 85.00 | 0.00 |
| 2025-11-19 | Wed | 15.0 | 20.8 | 7.7 | 50 | 10.08 | 7.54 | 62 / 91.00 | 0.00 |
| 2025-11-20 | Thu | 14.1 | 21.9 | 7.7 | 50 | 9.03 | 6.34 | 78 / 96.00 | 0.00 |
| 2025-11-21 | Fri | 13.4 | 24.8 | 7.7 | 50 | 5.88 | 3.87 | 78 / 97.00 | 0.00 |
| 2025-11-24 | Mon | 16.8 | 26.9 | 7.7 | 50 | 4.44 | 1.76 | 79 / 86.00 | 0.00 |

## Notes

### Competing Interest Statement

The authors have declared no competing interest.

### Funding Statement

This study was funded by
King's College equipment fund and Gates foundation

### Summary of Updates

1. In the results section, figures and table numbers cited in the text were revised and corrected. In some cases, there was a mismatch between the table numbers cited in the text and the real table numbers. 2. Headings of supplemental files were updated from Annex 1, 2.... to Supplemental Table 1 and 2. 3. Author affiliation institution for Gebeyehu Begashaw Abate added. The added institution is the University of Gondar, Ethiopia. Note: Except for such minor revisions, there are no significant differences between this version and the previous version.

